# Effect of electroMagnetic Interference from smartPHone On cardiac implaNtable Electronic device (EMI-PHONE study)

**DOI:** 10.1101/2021.11.05.21265956

**Authors:** Sanatcha Apakuppakul, Nilubon Methachittiphan, Sirin Apiyasawat

## Abstract

**Background:** Smartphone can emit two types of electromagnetic waves, static field from magnet and dynamic field from calling. Previous evidence showed the interference effects from old generation of mobile phones to cardiac implantable electronic device (CIEDs). The current generation of smartphones and CIEDS are reportedly better designed to reduce electromagnetic interference (EMI). We seek to find the presence and the magnitude of EMI from the current generation of smartphones.

**Methods:** A total of 80 consecutive subjects with CIEDs (Included pacemaker, ICD, CRT-D, CRT-P) were recruited from our CIEDs clinic and were tested for EMI. Each subject was tested with three different smartphones (Nokia 3310, Iphone 7, and Samsung 9S). Each phone was attached to chest wall at 0 cm at pulse generator site, at atrial lead, and at ventricular lead site. During the tests, real-time interrogations were performed to detect any EMI from smartphone in stand-by mode, and during calling-in and out for 30 seconds. After the tests, post-test interrogation was performed to detect any parameters changes. Adverse events including pacemaker inhibition, false ICD shock, CIEDs device malfunction, and urgent electrophysiologist consultations were recorded.

**Results:** Of all 80 subjects (Mean age 70.5±12.9 year-old, 50% male) recruited in the study, all completed the tests according to our protocol. The most common type of CIEDs tested was pacemaker (N=56, 70%), followed by ICD (N=16, 20%), and CRT (N=8, 10%). Most patients (N=62, 77.5%) had more than one lead implanted. The mean year of implantation was 5.2±2.8 (Devices were implanted since 2008-2019). Of all the tests performed, there was no EMI or adverse events observed.

**Conclusion:** Current generation of smartphones have no EMI effect to CIEDs and can be used safely without any adverse events including pacemaker inhibition, false ICD shock and CIEDs device malfunction.

## Introduction

Nowadays, smartphones are growth rapidly both number of users and technology. From national database of Thailand in 2018, 51 million smartphones were registered.

Smartphone can emit two types of electromagnetic waves, static field from magnet and dynamic field from calling. Previous evidence showed the interference effects from old generation of mobile phones to cardiac implantable electronic device (CIEDs).^1,2^

However, electromagnetic interference (EMI) from smartphones can effect cardiac implantable electronic devices (CIEDs) function by making noise signals and make CIEDs oversensing signals, leading to pacing inhibition in pacemaker and false shock in implantable cardioverter-defibrillator (ICD).^3.4^

There is recommendation from device companies and United States of America food and drug administration (US FDA) in using smartphones that mobile phones should be used at least 15 centimeters from devices.^5^

However this recommendation was derived from older generation of mobile phones and older generation of cardiac implantable electronic devices (CIEDs). The current generation of smartphones and CIEDS are reportedly better designed to reduce electromagnetic interference (EMI) and detect electromagnetic interference (EMI). The modern smartphone technology utilizing 4G or 5G frequency spectrum emits less electromagnetic waves than that of 2G or 3G technology.

There were a few studies that showed the safety in using smartphones less than 15 centimeters distance, but the number of subjects is small^6^ and limited mobile phones were tested. ^7^

We seek to find the presence and the magnitude of EMI from the current generation of smartphones.

## Methods

### Study population

Patient with cardiac implantable electronic devices (CIEDs) were recruited from Ramathibodi device clinic. Patients aged between 18 and 80 years old with any types of CIEDs were included in our study. Exclusion criteria are any abnormal CIEDs parameter during regular schedule device interrogation, for example, abnormal threshold, abnormal impedance, any leads and pulse generator issues. Pacemaker-dependent patients were excluded for safety reasons. During test protocol patient was closely monitored with cardiology fellow and device technician. Informed consent was obtained from every patient.

We initially planned to recruit at least 76 subjects in order to detect endpoints using 27% for effect of electromagnetic interference from previous study^3^ and 10% different from expected event with our study. Finally, we recruited 80 subjects since September 2018 – December 2019.

### Data collection and protocol flow chart

After patients visits Ramathibodi device clinic at regular follow-up schedule and devices parameter were in good ranges. The patients were informed consent and follow protocol testing as shown in figure 1. Baseline characteristic including age, sex, functional class, device type, mode of device, indication of implantation, position of implantation, number of leads, duration of implantation, LVEF and underlying heart disease, were recorded.

**Figure 1.**
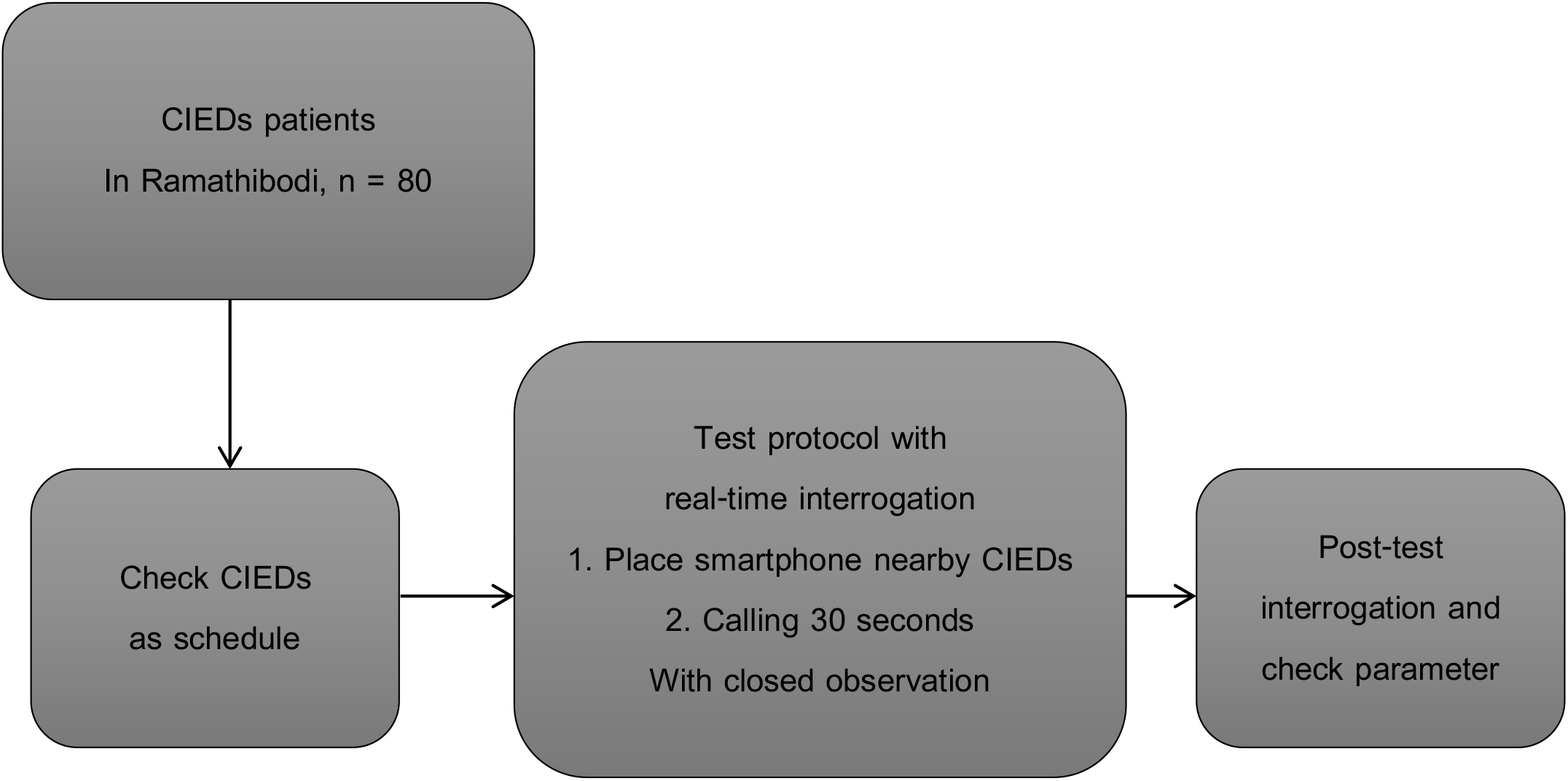
Protocol flow chart.

Smartphones were applied at 0 centimeter distance from patient’s chest wall. The position of smartphones were changed every 30 seconds during the test. These positions included pulse generator position, right parasternal border(right atrial lead site, if present), left parasternal border(right ventricular lead site) and apical area(left ventricular lead site, if present). Smartphones were tested in standby mode, 30-second calling-in and calling-out.

Smartphone brands used in our study are Nokia 3310, Iphone 7, and Samsung 9S. Each patient was tested with all 3 smartphones in all 3 modes (i.e. Standby, calling-in, and calling-out) with each smartphone placing at different positions as stated above. During the test protocol, real-time interrogation of device was done for detection of electromagnetic interference (EMI), pacemaker inhibition, false ICD shock, patients were closed monitoring with cardiology fellow and device technician stand-by. All real-time interrogated signal were printed for record. After protocol testing, device interrogation was done in all subjects to detect for any parameter changes from baseline. Testing protocol infographic was illustrated in figure 2.

**Figure 2.**
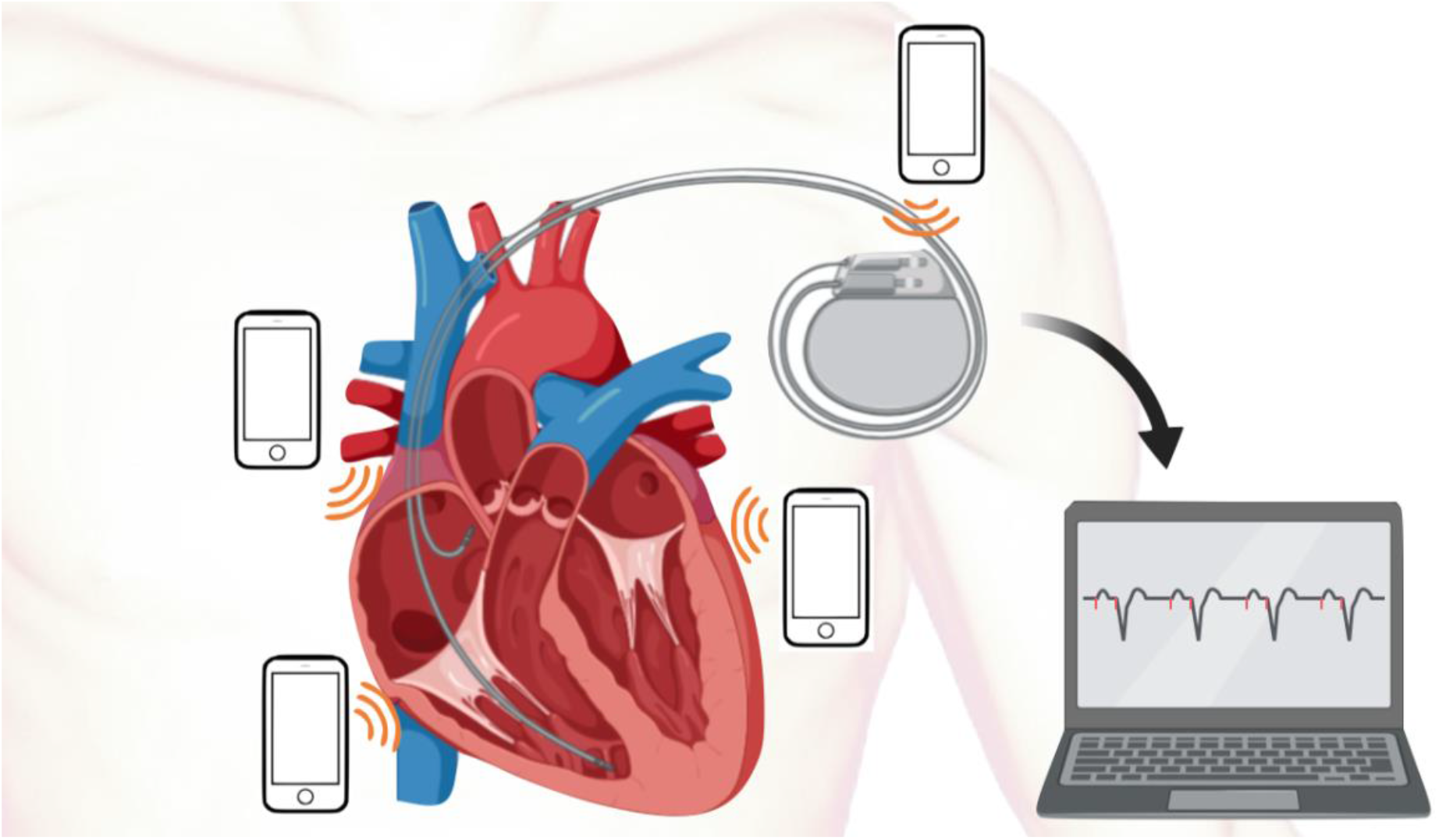
Infographic of testing protocol.

### Statistical analysis

Baseline characteristic are described as mean±SD for continuous data and proportions for categorical data. All CIEDs parameter are analyzed for detection of difference between pre- and post-test protocol, using t-test analysis. P-value less than 0.05 was used to detection of statistical significant. We use SPSS version 23 for data analysis.

### Ethical issues

All patient identifications were concealed prior to analysis, using number of case record form. Ethic approval to data collection and analysis were approved by the institutional review boards committees of Mahidol university. This study was complied with international guidelines for human research protection, Declaration of Helsinki, the Belmont report, CIOMS guidelines and the international conference on harmonization in good clinical practice (ICH-GCP). This study was funded by Ramathibodi research foundation after approval by ethic committee.

### Outcomes measurement

In our study, primary outcome is electromagnetic interference detected by real-time device interrogation. Every intracardiac electrogram was adjudicated with device technician, if any suspicion of EMI, adjudicated with electrophysiology fellow was performed. For secondary outcome are device malfunction detected post-protocol interrogation, pacing inhibition, false ICD shock, urgent electrophysiologist consultation, CCU admission.

## Results

### Baseline characteristics

A total 240 tests were performed in 80 enrolled subjects. Of all 80 subjects enrolled in our study, 40(50%) of patients were male. Mean age was 70.5 years old. NYHA functional class I and II were 92.5% and 7.5% respectively. Most of CIEDs device were pacemaker 56(70%) subjects, the others were ICD 16(20%) subjects, CRT-P 3(3.8%) subjects, and CRT-D 5(6.3%) subjects. Mode of device were DDD 52(65%) subjects, VVI 20(25%) subjects and biventricular pacing 8(10%) subjects. Majority of implanted device position was left pectoral region 74(92.5%) subjects. Patient who had 2-lead device were 53 subjects(66.3%), 1-lead were 18(22.5%) subjects and 3-lead were 9(11.3%) subjects. Mean duration of implantation was 5.2±2.8 years. Mean LVEF was 56.4±16.9%. Indication of implantation and underlying cardiac disease were described in table 1. Model of device was described in table 2.

**Table 1.** Baseline characteristics.

| Baseline characteristics | N (%) |
| --- | --- |
| Age (Mean±SD) | 70.5±12.9 year-old |
| Sex (Male) | 40(50%) |
| Functional class (NYHA) |  |
| NYHA class I | 74(92.5%) |
| NYHA class II | 6(7.5%) |
| Device type |  |
| Pacemaker | 56(70%) |
| ICD | 16(20%) |
| CRT-P | 3(3.8%) |
| CRT-D | 5(6.3%) |
| Mode of device |  |
| VVI(R) | 20(25%) |
| DDD(R) | 52(65%) |
| BiV | 8(10%) |
| Left pectoral implantation | 74(92.5%) |
| Number of leads |  |
| 1 lead | 18(22.5%) |
| 2 leads | 53(66.3%) |
| 3 leads | 9(11.3%) |
| Duration of implantation | 5.2±2.8 years |
| Years of implantation |  |
| 2008-2010 | 8(10%) |
| 2011-2013 | 20(25%) |
| 2014-2016 | 24(30%) |
| 2017-2019 | 28(35%) |
| LVEF (Mean±SD) | 56.4±16.9% |
| LVEF≤ 40% | 18(22.7%) |
| Underlying cardiac disease |  |
| Sinus node dysfunction | 21(26.5%) |
| Coronary artery disease |  |
| **Valvular heart disease** | 16(20%) |
| Severe AS |  |
| Severe MR | 3(3.9%) |
| Severe PR | 4(5.1%) |
| Severe TR | 1(1.3%) |
| **Cardiomyopathy** | 1(1.3%) |
| DCM |  |
| ICM | 14(17.5%) |
| HCM | 11(13.8%) |
| ARVC | 3(3.8%) |
| **Congenital heart disease** | 2(2.5%) |
| ASD |  |
| TOF | 1(1.3%) |
|  | 1(1.3%) |
| **Indication of device implantation** |  |
| Sinus node dysfunction | 25(31.5%) |
| AV block | 33(41%) |
| SCD: Primary prevention | 14(17.5%) |
| SCD: Secondary prevention | 8(10%) |

**Table 2.** Device model.

| Device model | N (%) |
| --- | --- |
| **Boston Scientific** |  |
| INOGEN MINI(D002) | 1(1.3%) |
| INOGEN EL(D140, D141) | 7(8.8%) |
| TELIGEN(F102, F103) | 2(2.5%) |
| AUTOGEN(F140, G058) | 4(5%) |
| LATITUDE(F141, F143) | 2(2.5%) |
| RESONATE(G447) | 1(1.3%) |
| CONTAK RENEWAL TR2(H145) | 1(1.3%) |
| INGENIO(J063, J065, J066, J177) | 10(12.5%) |
| ALTRUA(S501, S502, S503) | 9(11.3%) |
| ESSENTIO(L110, L111) | 22(27.5%) |
| VISIONIST(U228) | 1(1.3%) |
| **Medtronic** |  |
| ADAPTA DR(ADDR01) | 1(1.3%) |
| CONSULTA CRT-P(C3TR01) | 1(1.3%) |
| VIVA S CRT-D(DTBB2D1) | 2(2.5%) |
| EVERA MRI S(DVMC3D1, DDMC3D4) | 2(2.5%) |
| ENSURA MRI(EN1DR01) | 6(7.5%) |
| SENSIA D(SED01, SEDR01) | 4(5%) |
| VERSA DR(VEDR01) | 4(5.0%) |

### Outcomes

Total of 240 tests were performed on 80 subjects, no electromagnetic interference detection, no pacing inhibition, no false ICD shock was detected by real-time interrogation of CIEDs during standby mode, 30-second calling-in and 30-second calling out at any position and any brand of smartphones. No urgent EP consultation, no CCU admission was detected in our study, event rate was described in table 3. No any device parameter was significantly changed, including pacing function, sensing function, impedance and threshold, before and after out test protocol, device parameter was described in table 4.

**Table 3.** Event rate.

| Outcomes | Event(%) |
| --- | --- |
| **Primary outcome:** |  |
| Electromagnetic interference detection | 0 |
| **Secondary outcomes:** |  |
| Pacing inhibition | 0 |
| False ICD shock | 0 |
| Urgent EP consultation | 0 |
| Significant parameter change | 0 |
| CCU admission | 0 |

**Table 4.** Device parameter, pre- and post-test protocol.

| Parameter | Pre-test(Mean±SD) | Post-test(Mean±SD) | p-value |
| --- | --- | --- | --- |
| RA pace | 21.25±30 % | 22.5±30.6 % | 0.79 |
| RA sense | 2.4±2.1 mV | 2.4±2.2 mV | 1.0 |
| RA impedance | 353±220 Ohms | 359±221 Ohms | 0.86 |
| RA threshold | 0.46±0.4 mV | 0.45±0.4 mV | 0.88 |
| RV pace | 56±46 % | 56.1±46.1% | 0.99 |
| RV sense | 8.2±7.4 mV | 8.6±7.8 mV | 0.74 |
| RV impedance | 490±101 Ohms | 480.1±100 Ohms | 0.53 |
| RV threshold | 0.9±0.3 mV | 0.9±0.4 mV | 1.0 |
| LV pace | 97.8±2.9 % | 97±3.5 % | 0.74 |
| LV sense | 16.4±6.5 mV | 17.2±7.1 mV | 0.89 |
| LV impedance | 779±328 Ohms | 803±280 Ohms | 0.88 |
| LV threshold | 1.6±1.1 mV | 1.58±0.98 mV | 0.97 |
| Shock impedance | 49.3±10.2 Ohms | 50±10 Ohms | 0.83 |

## Discussion

The presence of electromagnetic interference (EMI) from older generation smartphones can effect cardiac implantable electronic devices (CIEDs) function by emission of noise signals. Older generation of CIEDs can be oversensing signals, leading to pacing inhibition in pacemaker and false shock in implantable cardioverter-defibrillator (ICD). Werner I. et. al. was described electromagnetic interference of pacemakers by mobile phones.^3^ Chiladakis et. al. was described in-vivo testing of digital cellular telephones in patients with implantable cardioverter-defibrillators.^4^ However, in contemporary era, newer generation of CIEDs and smartphones were rapidly improved in technology. CIEDs had many new features, including noise detection mode. Smartphones were also improved in network signaling, including 3G, 4G and 5G in present. Burri H et. al. was published low risk of electromagnetic interference between smartphones and contemporary implantable cardioverter defibrillators.^6^ Lennerz C et. al. was revealed safety of using smartphones on CIEDs in electromagnetic interference aspect.^7^

In our study, we established safety of using current generation smartphones and CIEDs.

We revealed that no any EMI was detected by real-time interrogation of CIEDs in our test protocol, no any parameter changes.

From this study, we demonstrated safety and extremely low-risk of effect of EMI on CIEDs and proposed that current recommendation should be changed and current generation of smartphones can be used safely within 15 cm from pulse generator and leads.

## Limitations

In our study, we have some limitation, first we lacked of comparison group. Second, we have no electromagnetic machine for detect true value of electromagnetic wave. Third, we have only two device companies (Boston scientific, Medtronic), three mobile phones model (Nokia 3310, Iphone 7, and Samsung 9S) and 3G, 4G systems were tested in our study.

## Conclusion

Current generation of smartphones have no EMI effect to CIEDs and can be used safely without any adverse events including pacemaker inhibition, false ICD shock and CIEDs device malfunction. We suggested that current recommendation should be changed and current generation of smartphones can be used safely within 15 cm from pulse generator and leads.

## Data Availability

All data produced in the present work are contained in the manuscript

---

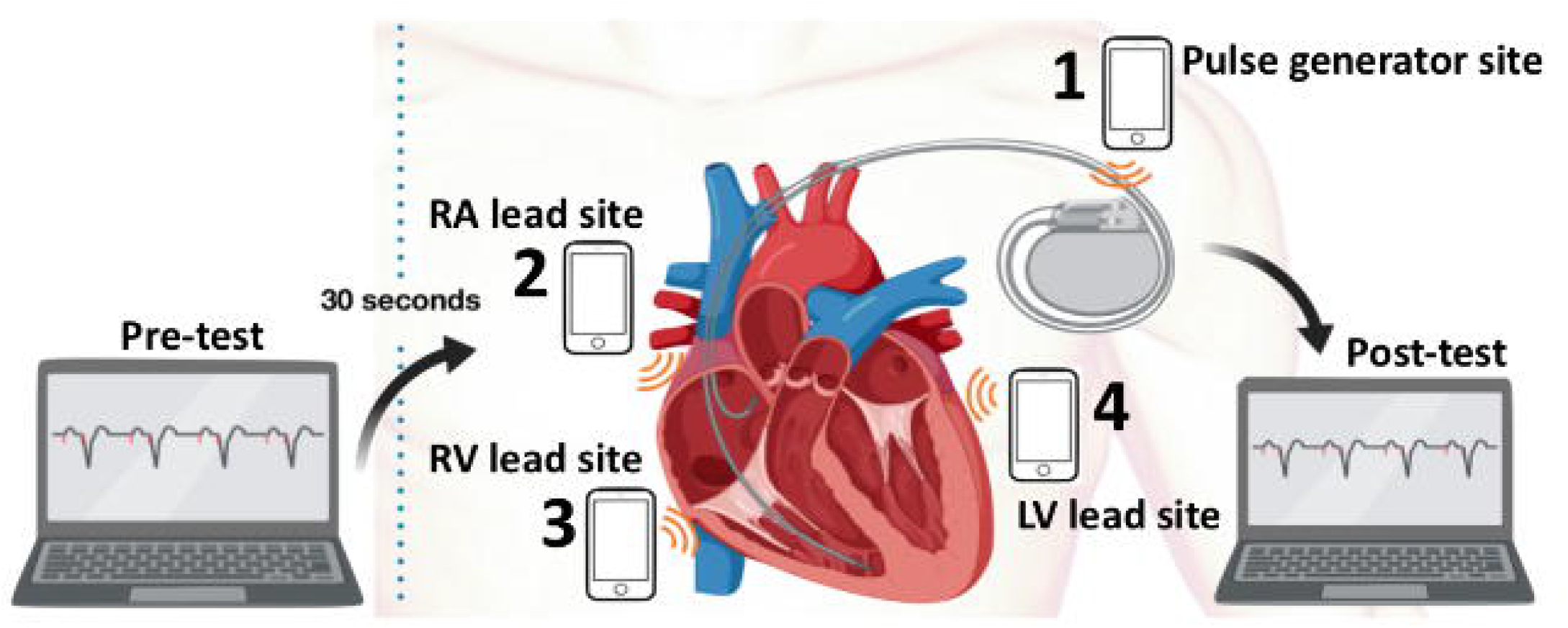

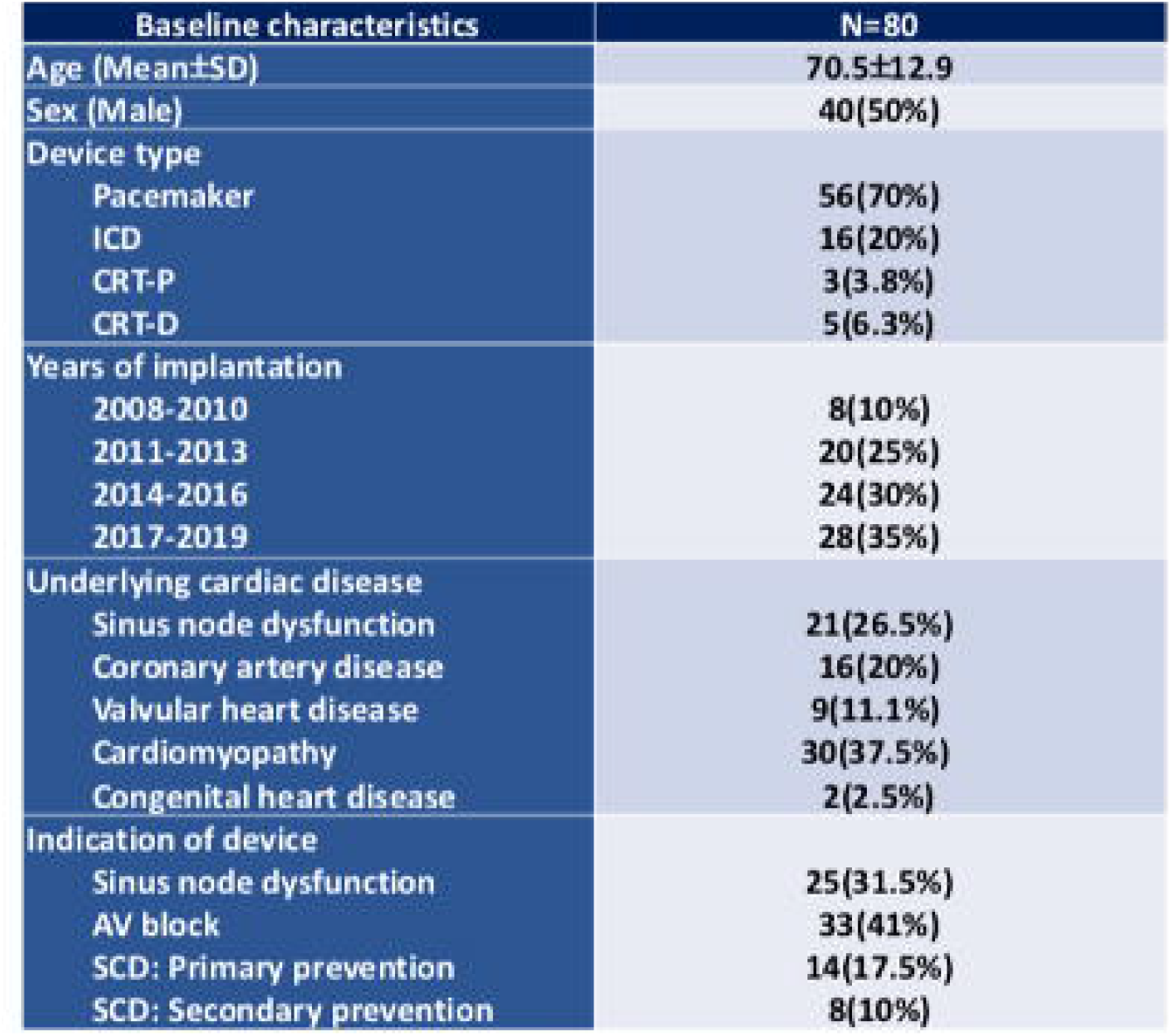

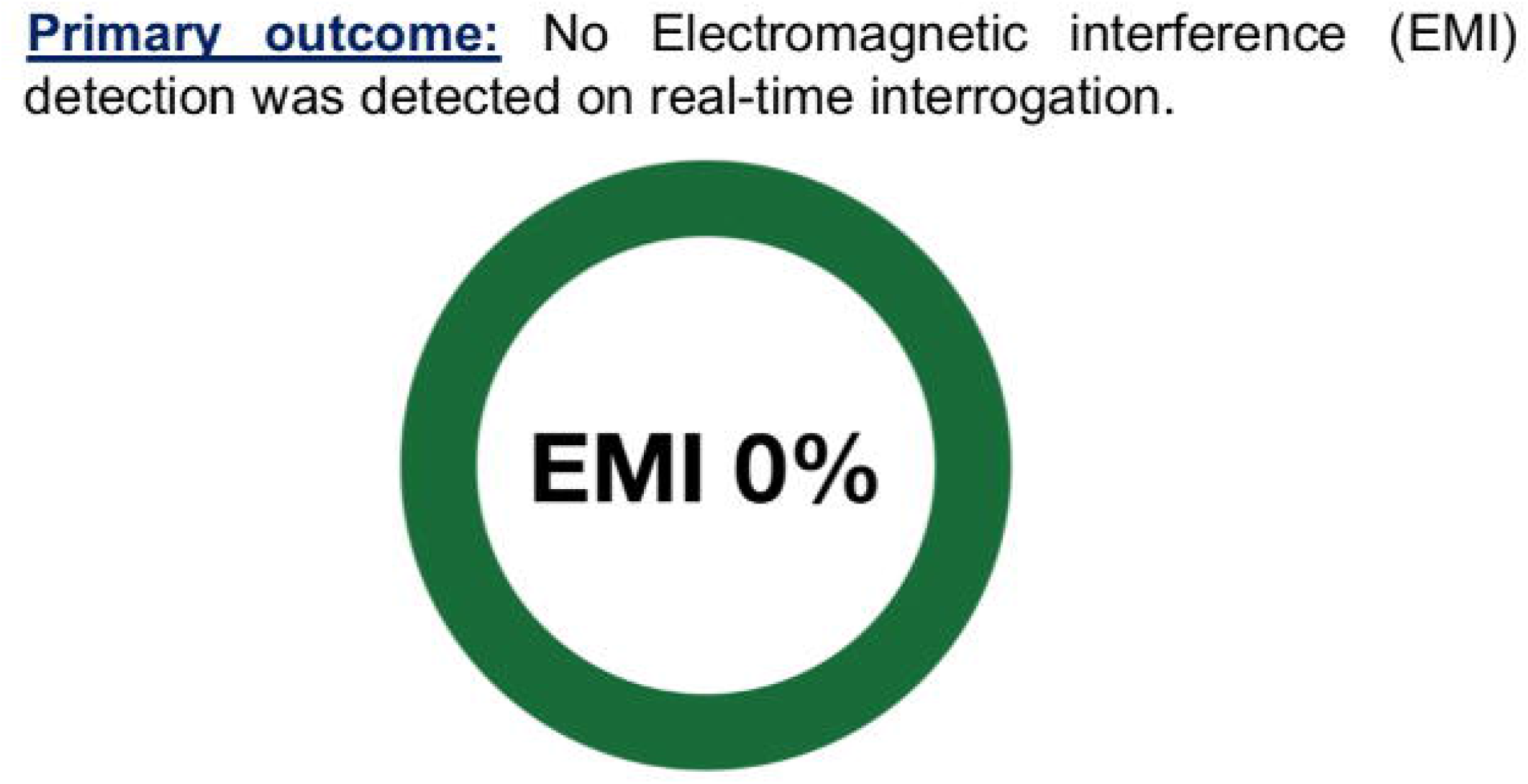

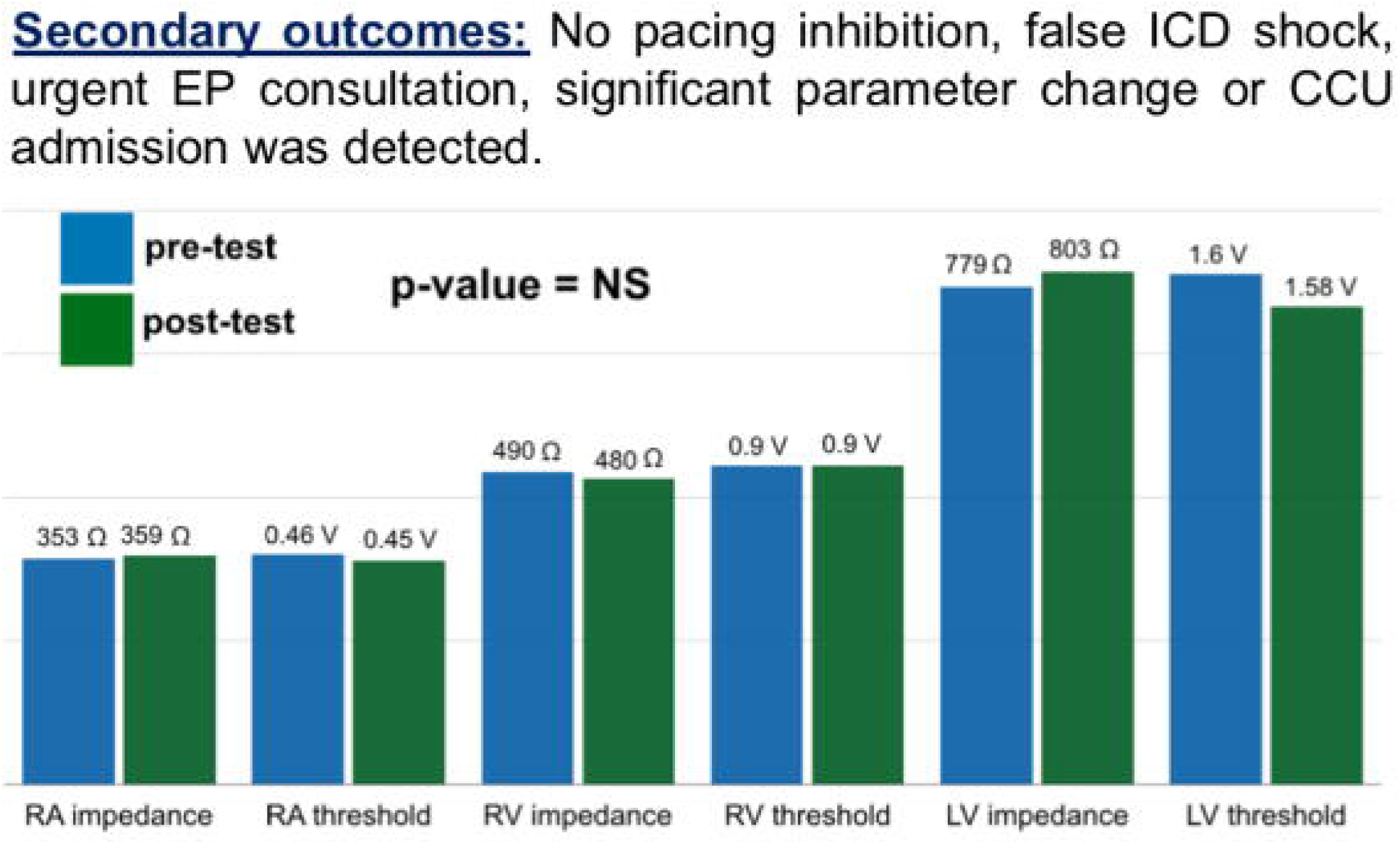

